# Identification and Attribution of Weekly Periodic Biases in Epidemiological Time Series Data

**DOI:** 10.1101/2023.06.13.23290903

**Authors:** Kit Gallagher, Richard Creswell, David Gavaghan, Ben Lambert

## Abstract

COVID-19 data exhibit various biases, not least a significant weekly periodic oscillation observed globally in case and death data. There has been significant debate over whether this may be attributed to weekly socialising and working patterns, or is due to underlying biases in the reporting process. We characterise the weekly biases globally and demonstrate that equivalent biases also occur in the current cholera outbreak in Haiti. By comparing published COVID-19 time series to retrospective datasets from the United Kingdom (UK) that are not subject to the same reporting biases, we demonstrate that this dataset does not contain any weekly periodicity, and hence the weekly trends observed both in the UK and globally may be fully explained by biases in the testing and reporting processes. These conclusions play an important role in forecasting healthcare demand and determining suitable interventions for future infectious disease outbreaks.

## 1 Introduction

A significant periodic oscillation has been identified in US COVID-19 time series datasets for both case and death data [1], which has since been substantiated by observations of Huang et al. [2] on worldwide COVID-19 data. While, to the best of our knowledge, such variation has not been reported for other diseases, our results in this paper indicate that a similar weekly periodic trend exists in an ongoing multi-country outbreak of cholera. Identifying the cause of variation in reported epidemiological data (and specifically whether there is a genuine underlying trend in epidemiological events or whether this is simply an artefact of the reporting process) is crucial for forecasting healthcare demands such as numbers of intensive care beds.

Having declared COVID-19 to be a global pandemic, the World Health Organization (WHO) suggested that early detection and prompt response would be critical in slowing down the spread of the the outbreak. The collection and prompt publication of datasets recording occurrences of cases and deaths has been crucial to this response in regions across the world. In contrast, the response to the ongoing resurgence of Cholera (with over 14,000 suspected cases in Haiti) has been limited by our degree of understanding of current disease dynamics. A recent (22nd March) WHO report [3] on the ongoing multi-country outbreak of cholera classified the global risk as “Very High”, reporting that strengthened surveillance and timely case management are urgently needed.

Here, we will first characterise these periodic reporting biases within the cholera and COVID-19 datasets, considering both case and death data for COVID-19 globally. We then discuss the origin of these data, before finally using a unique dataset from the United Kingdom (UK) to demonstrate that such periodic trends in COVID-19 may be fully attributable to biases in national reporting processes.

## 2 Methods and Data

The COVID-19 data used in this report were extracted from the John Hopkins Database [4], up to 1st March 2023, and correspond to the UK unless otherwise stated. Case rates are based on the total number of positive tests (accounting for individuals taking multiple tests), while death rates are defined as the number of deaths with a positive COVID-19 test in the past 28 days. The raw data give daily cumulative totals for both cases and deaths, which we used to generate daily incidences and daily deaths. Any negative counts (resulting from a decrease in the cumulative total) were attributed to changes in the reporting mechanism, and therefore excluded from further analysis.

Case incidence time series of the cholera outbreak in Haiti were obtained from the PAHO/WHO Cholera dashboard up to 4th April 2023, available at: https://shiny.pahobra.org/cholera/. This combines case reports from all 10 departments of Haiti reported by the Haiti Ministry of Public Health and Population.

To characterise the periodic biases observed in both datasets, we define a reporting factor *α*_*i*_ for each day *i*, given by the ratio between the observed value on a given day and the average value from a seven-day window centred on the given day. Global trends in this weekly bias for COVID-19 were also characterised using Principal Component Analysis (PCA) from over 200 countries included in the John Hopkins database. We have additionally developed a user-friendly, open-source Python library for exploring and visualizing periodic trends in COVID-19 data, which includes notebooks for reproducing all results appearing in this paper, available at: https://github.com/KCGallagher/periodic-sampling.

## 3 Results

### 3.1 Weekly Reporting Trends - COVID-19

Figure 1a shows the distribution of the reporting factors, *α*_*i*_, globally for COVID-19 time series data. Significant under-reporting over the weekend observed for most countries can be clearly discerned.

**Figure 1:**
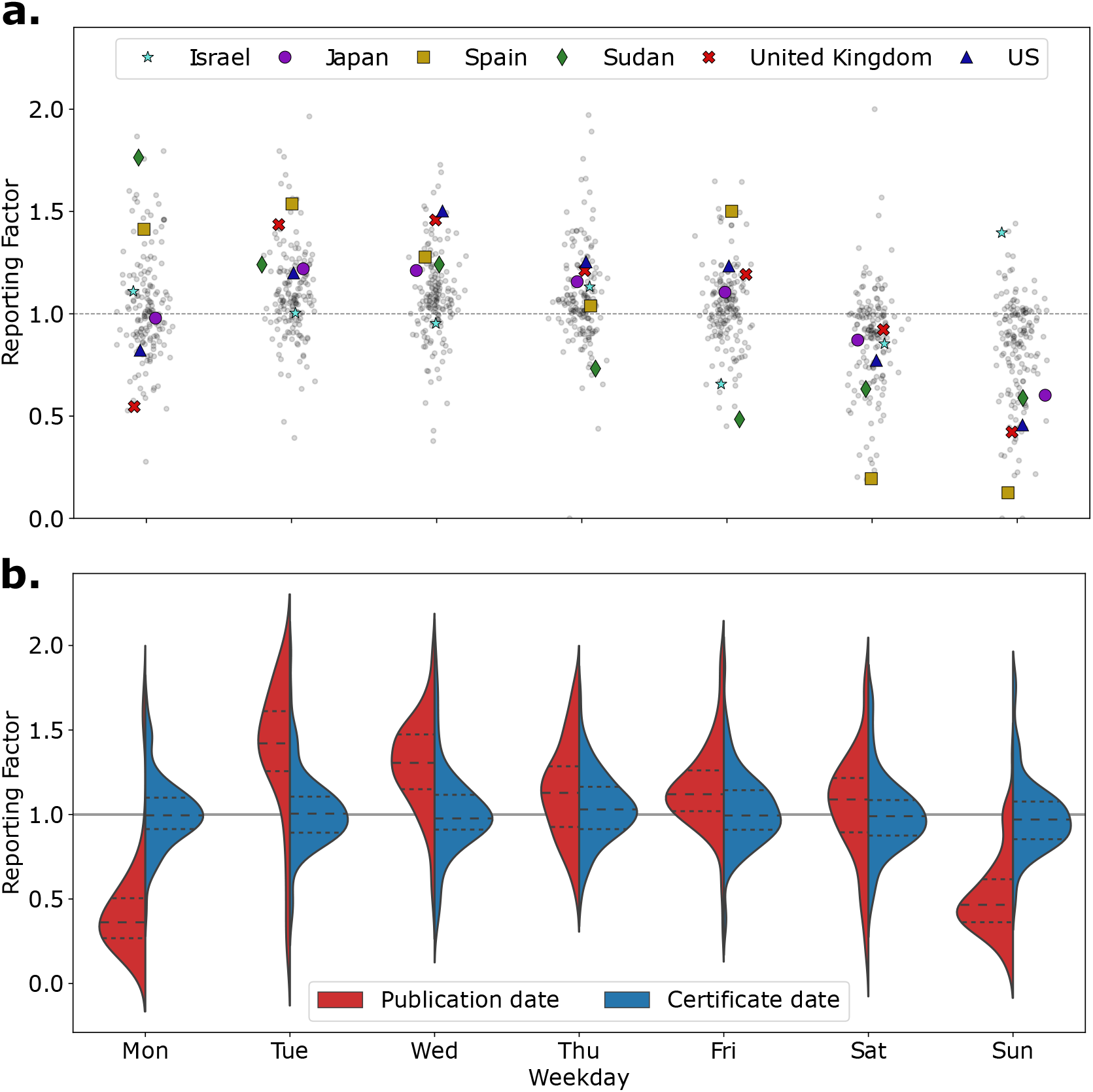
**(a)** Distribution of reporting factor values (grouped by weekday) for daily death statistics globally, with selected countries highlighted. (Gaussian jitter was applied to x-axis values for visualisation purposes). **(b)** Distribution of reporting factors in UK death data, with interquartile ranges marked as horizontal dashed lines. A strong bias is observed in the death data grouped by publication date, which does not occur when deaths are attributed to the day marked on the death certificate.

Table 1 gives the two primary principal components from the PCA; it is clear that the largest principal component (accounting for over a third of the variation across the dataset) corresponds to weekend under-reporting, which is broadly compensated for across the week as a whole. More interestingly, the second principal component corresponds to the under-reporting on Mondays; the ten countries with the highest PC2 scores only had an average reporting factor of 0.46 on Mondays. This trend was observed in both the US and UK data and appears to exist separately to weekend under-reporting. We do not have a hypothesis for the origin of these trends.

**Table 1:**
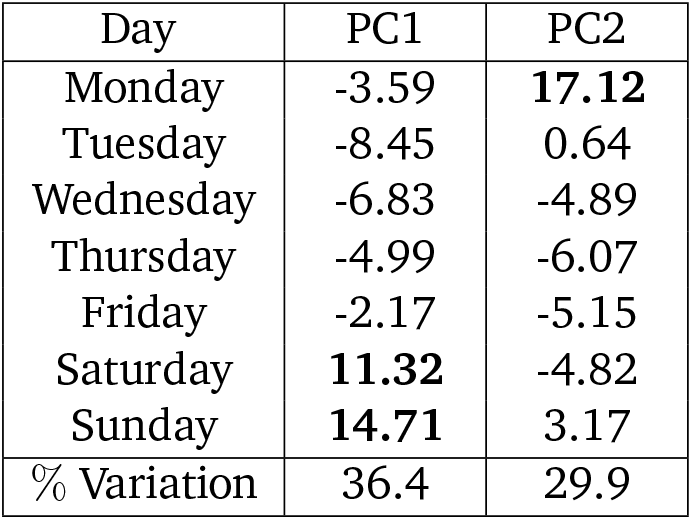
The first two principal component loadings across 200 countries, which illustrate the magnitude of reporting biases by day of week. The first component demonstrates clear weekend under-reporting being compensated for during the week, while the second component suggests that under-reporting on Mondays may also be present for a set of countries.

| Day | PC1 | PC2 |
| --- | --- | --- |
| Monday | -3.59 | **17.12** |
| Tuesday | -8.45 | 0.64 |
| Wednesday | -6.83 | -4.89 |
| Thursday | -4.99 | -6.07 |
| Friday | -2.17 | -5.15 |
| Saturday | **11.32** | -4.82 |
| Sunday | **14.71** | 3.17 |
| % Variation | 36.4 | 29.9 |

### 3.2 Weekly Reporting Trends - Cholera

Applying these periodic analysis methods instead to daily case data from Haiti, we also observed consistent weekly trends characterised by a significant under-reporting on Sundays. Computing the Kruskal-Wallis H statistic on the reporting factor distribution between weekdays, we found significant variation (*H* = 356, *DF* = 5, *p <* 0.01) in the reporting factor distribution between weekdays.

### 3.3 Origin of Periodic Bias

Bergman et al. [1] attribute periodic oscillation in COVID-19 data to weekly fluctuations in case reporting (such as a consistent under-reporting bias at weekends compensated by over-reporting during the week), a hypothesis supported by the analysis from Hotz et al. of case incidence data in Germany [5]. This association is disputed, however, and it has been suggested that these fluctuations reflect an underlying periodicity in the true case/death data, either due to weekly variation in inter-generational interactions [6], working patterns [7], or even an underlying circadian rhythm of the virus [8].

Biases in the reporting process may be identified by distinguishing between the data published by date recorded or date reported. While the data by date recorded ultimately provide the most accurate representation of the pandemic, these are only available in a handful of countries/regions, and are typically significantly delayed to ensure that all historic events are included. Decision–making agencies therefore rely on data by date reported, which at a given point in time represent the current best understanding of the pandemic, but are hampered by delays in reporting [9].

While global data typically group both cases and deaths by date reported (hereafter referred to as ‘publish date’), data from the UK government website offer a unique opportunity to identify biases in the reporting process, distinguishing between deaths grouped by date published and those grouped by date recorded on the death certificate. While such data are not immune from reporting delays, only being updated days or even weeks after the date in question, they do eliminate systematic, periodic biases, thereby allowing us to determine the role of reporting in generating the periodic oscillations observed in datasets grouped by publication date.

Computing the Kruskal-Wallis H statistic on the reporting factor distribution between weekdays for both COVID-19 death datasets, we found no evidence for weekly periodicity in the grouping by death certificate date (*H* = 6, *DF* = 5, *p* = 0.42), while there was strong variation (*H* = 356, *DF* = 5, *p <* 0.01) in the median reporting factor for the grouping by publication date. The differing distribution of daily reporting factors for each dataset is shown in Figure 1b, illustrating the weekly bias introduced by the publication process.

### 3.4 Source of Reporting Bias

Bukhari et al. [10] hypothesised that under-reporting was a result of increased strain on health services coinciding with reduced reporting capacity at weekends. We considered whether there was a linear relationship between the reporting factor and current pandemic size, assuming that the strain on healthcare would be well represented by the number of current cases (incidence count). In our regression, the slope coefficient on incidence count did not differ significantly from zero (*t* = 0.6, *DF* = 85, *p >* 0.05), failing to support the conclusion that there is a significant relationship between reporting factor and current incidence level of the pandemic.

## 4 Discussion

We have demonstrated that there is substantial periodic bias in COVID-19 datasets both from the UK and across the globe, with a clear weekly frequency. In any subsequent analyses, this bias is typically removed with a rolling seven-day average which inevitably causes a delay in the appearance of trends within the data, impacting the efficacy of government interventions. Despite sustained debate [6–8] on the cause of this oscillation, we have shown for the UK it can fully be explained by biases in the testing and reporting processes, and that datasets not subject to such biases do not exhibit any weekly periodicity.

We also demonstrate that similar periodic biases exist in Haitian data from the ongoing cholera epidemic. Given the water-based transmission mechanisms of this outbreak contrast dramatically with the primarily airborne transmission of coronavirus, it is likely such weekly trends are not isolated to these two outbreaks, instead being endemic across many epidemiological datasets. As our ability to track emerging epidemics in ‘real time’ increases due to improving temporal data resolution, it is increasingly important to develop knowledge and understanding of the biases in such datasets. These conclusions are highly relevant to healthcare providers in forecasting demand and to policymakers seeking to determine interventions for containing infectious disease outbreaks.

## Data Availability

All data used in the study are available at:
https://github.com/CSSEGISandData/COVID-19
https://shiny.pahobra.org/cholera/
All data analysis is available at:
https://github.com/KCGallagher/periodic-sampling

https://shiny.pahobra.org/cholera/

https://github.com/KCGallagher/periodic-sampling

## Author Contributions

All authors contributed to the study conception and design. Data collection and analysis were performed by Kit Gallagher. The first draft of the manuscript was written by Kit Gallagher and all authors commented on subsequent versions of the manuscript. All authors read and approved the final manuscript.

